# IMPACT OF GEOSPATIAL FOOD ACCESS ON ACUTE PANCREATITIS OUTCOMES

**DOI:** 10.1101/2024.02.27.24303446

**Authors:** Ankit Chhoda, Marco Noriega, Tamara Kahan, Anabel Liyen Cartelle, Kelsey Anderson, Shaharyar A. Zuberi, Miriam Olivares, Jill Kelly, Steven D. Freedman, Loren G. Rabinowitz, Sunil G. Sheth

## Abstract

**BACKGROUND AND AIM:** Food access is an important social determinant of health and refers to geographical and infrastructural aspects of food availability. Using publicly available data on food access from the United States Department of Agriculture (USDA), geospatial analyses can identify regions with variable food access, which may impact acute pancreatitis (AP), an acute inflammatory condition characterized by unpredictable outcomes and substantial mortality. This study aimed to investigate the association of clinical outcomes in patients with AP with geospatial food access.

**METHODS:** We examined AP-related hospitalizations at a tertiary center from January 2008 to December 2018. The physical addresses were geocoded through ArcGIS Pro2.7.0 (ESRI, Redlands, CA). USDA Food Access Research Atlas defined low food access as urban areas with 33% or more of the population residing over one mile from the nearest food source. Regression analyses enabled assessment of the association between AP outcomes and food access.

**RESULTS:** The study included 772 unique patients with AP residing in Massachusetts with 931 AP-related hospitalizations. One hundred and ninety-eight (25.6%) patients resided in census tracts with *normal* urban food access and 574 (74.4%) patients resided in tracts with *low* food access. AP severity per revised Atlanta classification [OR: 1.88 (95%CI: 1.21-2.92); *p*=0.005], and 30-day AP-related readmission [OR: 1.78(95%CI: 1.11-2.86); *p=*0.02] had significant association with food access, despite adjustment for demographics, healthcare behaviors, and comorbidities (Charlson Comorbidity Index). However, food access lacked significant association with AP-related mortality (*p*=0.40) and length of stay (LOS: *p=*0.99).

**CONCLUSION:** Low food access had a significant association with 30-day AP-related readmissions and AP severity. However, mortality and LOS lacked significant association with food access. The association between nutrition, lifestyle, and AP outcomes warrants further prospective investigation.

## INTRODUCTION

Food insecurity is defined by the Food and Agriculture Organization as uncertain access to safe, sufficient, and nutritious food, with influencing factors ranging from agricultural production and distribution systems to socioeconomic conditions.^1^ Individuals facing food insecurity are susceptible to increased risks of malnutrition, chronic diseases, and compromised physical and mental health.^2^ Food insecurity pertains to a household’s ability to afford and access enough food to meet its members’ nutritional needs and is frequently measured through patient-reported surveys. Factors contributing to food insecurity include income constraints, unemployment, poverty, and unexpected expenses. Existing evidence suggests that food insecurity may play a role in digestive health, and has been previously investigated in patients with liver diseases and cystic fibrosis.^3, 4^ Food insecurity overlaps with low food access, which refers to geographical and infrastructural aspects of food availability, such as the distance to the nearest food source. Access to nutritious food is an important social determinant of health and has an impact on health disparities and outcomes across diverse populations.^5, 6^

The ramifications of food access may extend to its impact on the development and/or recovery of acute pancreatitis (AP), a condition characterized by inflammation of the pancreas, which carries a substantial mortality rate of up to 25% in severe cases, and is associated with a complex recovery process.^7^ Annually, AP emerges as a notable contributor to gastrointestinal-related hospital admissions, accounting for 288,220 US hospitalizations in 2018. ^8^ The escalating burden of AP is evident in the annual percent change of 3.1%, indicating a rising trend.^9^ Improved clinical awareness of the condition, enhanced diagnostic techniques, and exposure to attributable risk factors such as alcohol use and smoking may contribute to this upward trajectory. ^10, 11^ The economic burden of AP is also substantial, with annual healthcare expenditures exceeding $2.5 billion in the United States (US).^12^ Through an exploration of the interconnected factors encompassing nutrition, lifestyle factors, and socio-economic conditions, investigation of the association between food access and the outcome of AP is warranted. While there is a paucity of literature linking AP outcomes to food access, this association can be hypothesized from extensive research on the health implications of the interplay between nutrition, lifestyle factors, and digestive health. Inadequate access to nutritious food may lead to malnutrition, which is a known risk factor for pancreatitis.^13^ A diet high in saturated fats may induce inflammation, which may also negatively impact the pancreas and exacerbate inflammation during AP.^14^ Socioeconomic factors like poverty, often linked with low food access, may coexist with additional pancreatitis risk factors, including alcohol abuse and smoking.^15^ Furthermore, stress-related modifications in the gut microbiota may also contribute to AP severity. ^16^ This observational study of patients with AP aims to examine the association between the severity and outcomes of AP with geospatial food access.

## METHODS

### Study Design and Setting

This single-center retrospective cohort study included patients diagnosed with AP and hospitalized at a tertiary care center in Massachusetts between January 1, 2008, and December 31, 2018. This study was approved by the Institutional Review Board (Protocol ID:2018P000613). This study is reported in accordance with the Strengthening the Reporting of Observational Studies in Epidemiology (STROBE) guidelines (**Supplement 1**).^17^

### Study Population

Adult patients with AP were identified through *International Classification of Diseases, Ninth and Tenth Revision* codes 577.1 & K85.9.^18, 19^ Electronic health records (EHRs) were then manually reviewed to confirm the diagnosis based on Revised Atlanta classification (2 of the following during hospitalization: typical abdominal pain, elevation of serum lipase level three times upper limit of normal, or evidence of pancreatitis on cross-sectional imaging).^20^ Patients who carried a diagnosis of chronic pancreatitis or pancreaticobiliary malignancy and those lacking a physical address, thus preventing geospatial coding, were excluded.

### Food Access Assessment

The physical addresses of AP patients hospitalized at a large, tertiary care center in Massachusetts between 1/2008-12/2018 were geocoded through ArcGIS Pro 2.7.0 (ESRI, Red-lands, CA). Using their census tracts, we determined physical food access using the geographic indicators from the US Department of Agriculture (USDA)’s Food Access Research Atlas, which estimate the accessibility of food sources from a list of supermarkets, the Decennial Census, and the 2014-2018 American Community Survey (ACS) (https://www.ers.usda.gov/data-products/food-access-research-atlas/go-to-the-atlas).^21^ Food access was calculated by the USDA based on the Euclidean distance from the centroid of each individual census tract to a nearby food sources such as a supermarket, bodega, or grocery store. The Census Bureau defines urban areas as geographical areas with a population of more than 2,500 people residing within a delimited census tract (**Supplement 2**). Thus, per USDA definitions, low food access was defined as residence in regions where 33 % or more of the population were more than one walkable mile from a food source.^21^ Normal food access was determined on all other census tracts with less than 33 % of the population residing more than one walkable mile from a previously defined food source. The physical food sources were extracted using the merged data from supplemental nutrition assistance program (previously “food stamp”) Supplemental Nutrition Assistance Program (SNAP) eligible stores in the STARS directory and the Trade Dimensions TDLinx directory of stores.^22, 23^

### Baseline Demographics and Covariates

Patient demographics (including age, sex, race and ethnicity, active smoking or alcohol use, etiology of AP, insurance type and address), and clinico-radiologic information were extracted through review of individual EHR.^24, 25^ The ethnoracial features were self-reported and verified by health care practioners. Additionally, we quantified the comorbidity burden for each participant using the Charlson Comorbidity Index (CCI).^26^

### Outcome Measures

1. <u>Primary outcome:</u> Severity of AP was based on the Revised Atlanta classification: mild AP (no local or systemic complications), moderately severe AP (transient organ failure, local complications, or exacerbation of comorbidities) and severe AP (characterized by persistent (>48 hours) organ failure).^20^ Local complications included pseudocysts, peripancreatic collection, pancreatic necrosis, and splenic vein thrombosis. Systemic complications included respiratory failure requiring intubation, renal failure, bacteremia, and sepsis.
2. <u>Secondary outcomes:</u> We also collected data on AP outcomes, including the length of hospital stay (LOS) for each patient, magnitude of pain upon presentation and discharge quantified by visual analog scale (VAS), opioid use quantified through morphine equivalent units (MME) required during the entire hospitalization. We recorded the disposition of patients, including discharges to extended care facilities, the number of readmissions within 30 days, and mortality within one year of original hospitalization. Furthermore, we collected data on extra-pancreatic complications such as gastrointestinal bleeding and delirium.

### Statistical Analysis

Categorical data were presented as proportions and continuous variables were reported as medians with interquartile range (IQR). Statistical differences in categorical variables were assessed using Chi-square/Fisher’s exact tests, while continuous variables were analyzed using Kruskal-Wallis tests. AP outcome variables were considered a dependent variable and underwent logistic regression analysis. When univariate regression analysis was performed, associations with a significance threshold of p-value < 0.1 were included in the multivariable regression models. The multivariable model was adjusted for baseline factors including age, ethnoracial distribution, sex, and comorbidities (quantified using CCI) and health care behavior. The statistical analysis was carried out using STATA (StataCorp LLC, Version 17.0, College Station, TX). A p-value ≤ 0.05 was considered statistically significant.

## RESULTS

### Study Population Characteristics and Food Access Assessment

The study included 772 patients residing within the state of Massachusetts with a median age of 53.1(11) years and 49.9% of patients were female. We noted 198(25.6%) patients residing in census tracts with <u>normal</u> urban food access and 574 patients (74.4%) in tracts with <u>low</u> food access. **(Figure 1A)**. The number of hospitalizations for AP for patients residing in a normal food access tract was 229 (24.6 %) and low food access was 702 (75.4%). In this study, patients predominantly self-identified as non-Hispanic White (n=463; 70.86%). Under-represented ethno-racial groups comprised 144 (21.73%) non-Hispanic Black, 101(1.25%) Hispanic, and 27 (3.5%) non-Hispanic Asian, 9(0.47%) Native American patients. Seven patients had mixed race whereas 21 patients lacked ethnoracial data. There was no statistical difference between body mass index (BMI) of patients with AP (median (IQR)) and those with normal and low food access (26.95(4.3) *vs*. 26.7 (4); *P=* 0.11). The majority of the patients relied on federal/state insurance and the difference between the low and normal food access groups lacked statistical significance. The etiology for AP was predominantly biliary and baseline demographics have been summarized in **Table 1**.

**Table 1:**
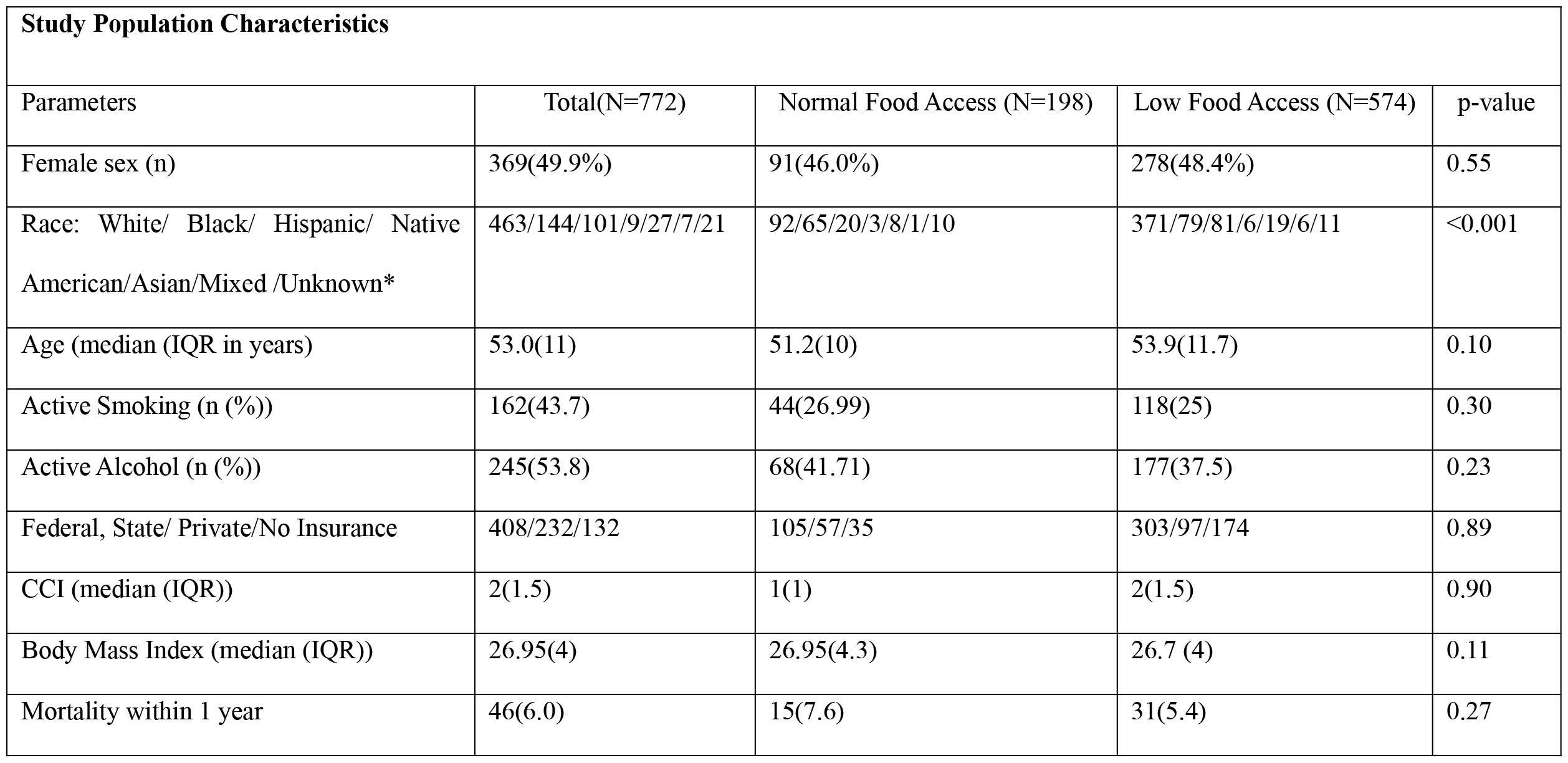
Description of baseline demographic features of acute pancreatitis patients.

**Figure 1(A):**
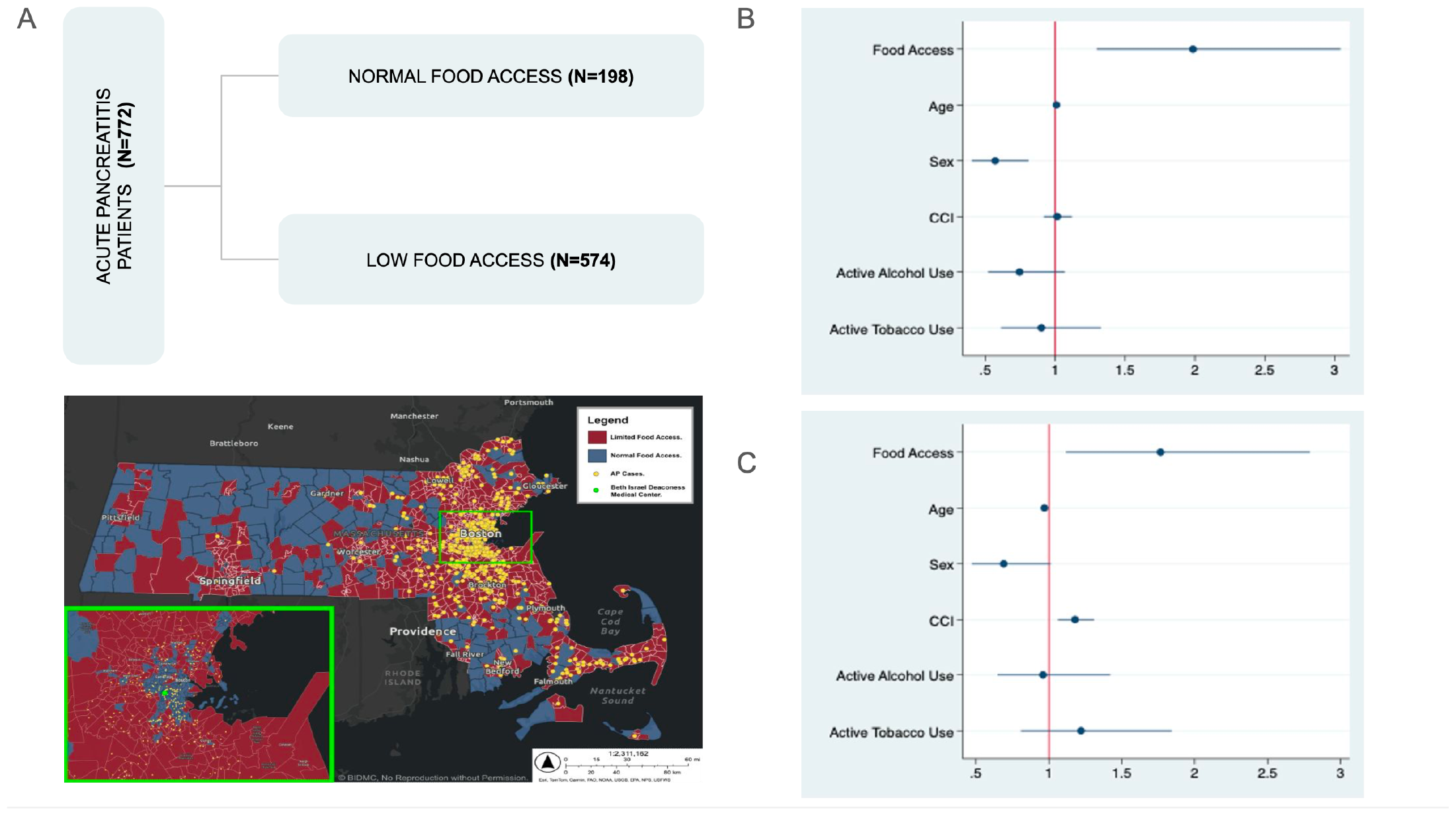
**Acute Pancreatitis Cohort Food Access Distribution in Massachusetts. Association of Geospatial Food Access after adjusting for demographic variables (age, CCI, ethnoracial distribution, sex) & health care behaviors (active alcohol and tobacco use) with (B) AP severity and (C) 30-day readmission rate.**

### Clinical Course and Management

1. <u>AP severity:</u> A significantly higher prevalence of moderately severe [133/702 episodes of AP (18.9%)] and severe AP [140/702(20.1%)] was observed among patients with low food access as compared to normal food access [30/229 episodes of AP(13.1%) and 38/229(16.6%) respectively, *p=* 0.03]. The occurrence of AP-related systemic complications, including respiratory failure, acute kidney injury, bacteremia and sepsis did not show statistical significance across the food access groups. The BISAP score (median (IQR)) of patients with low geospatial food access was higher as compared to those with normal access [1(1) vs. 1(0.5), *p=* 0.02]. The clinical and disposition outcomes of patients with AP have been summarized in **Table 2**.
2. <u>Management:</u> The magnitude of pain quantified by VAS lacked significant association in the groups residing in low vs. normal food access at presentation [7(2) and 8(1.5) (median (IQR), respectively, *p=*0.53] and upon discharge [0(1) and 0(0) respectively, *p=* 0.46]. Their opioid use quantified by MME requirements during the hospital stay lacked statistical significance [normal food access: 16(17) *vs*. low food access: 12.8(30); *p=* 0.42]. Early enteral feeding was initiated at 2(1) days of presentation in the overall AP population and was comparable in low and normal food access residential regions [1(1) and 2(1) respectively, *p=* 0.54].

### Disposition outcome

Within one year of follow-up, 46 patients died, and we found no significant difference in mortality based on residence, comparing normal food access [15 (7.6%)] to low food access [31 (5.4%), p=0.27]. Additionally, we also observed 125(16.2) AP-related readmissions within 30 days of discharge. Notably, regions with low food access showed a higher rate of 30-day AP-related readmissions compared to those with normal access [135/702 (19.2%) vs. 31/229 (13.5%); p=0.04]. Majority of the patients with AP were discharged to home and we noted 27 (3%) patients requiring rehabilitation center placement which did not differ between patients with normal vs. low food access (p= 0.60).

**Table 2:** Outcomes in patients with acute pancreatitis-related hospitalizations (N= 931)

| AP-related hospitalization outcomes |  |  |  |  |
| --- | --- | --- | --- | --- |
| Parameter | Total(N=931) | Normal Food Access (n=229) | Low Food Access(n=702) | p-value |
| Severity (Modified Atlanta Criteria) | Mild: 590(63.3)<br>Moderate:163(17.5)<br>Severe: 178(19.2) | Mild:161(70.3%)<br>Moderate:30(13.1%)<br>Severe: 38(16.6%) | Mild:429(61.0%)<br>Moderate:133(18.9%)<br>Severe: 140(20.1%) | 0.03 |
| <u>Etiology:</u> Biliary/Alcohol/PEP/HyperTG/<br>Idiopathic/Mass/ Other/ Genetic/ Drugs | 286/197/20/17/141/6<br>7/170/21/12 | 72/51/5/4/39/13/39/5/4 | 214/146/15/13/102/54/1<br>31/16/8 | 0.20 |
| 30-day rehospitalization [n (%)] | 159(17.8) | 28(13.5) | 131(19.2) | 0.04 |
| BISAP (median (IQR)) | 1(1) | 1(0.5) | 1(1) | 0.02 |
| Length of stay [median (IQR)] | 4(2.5) | 4.0(2.3) | 4.2(2.6) | 0.92 |
| IV Analgesia (Yes/No/Unknown) | 645/75/30 | 479/60/26 | 166/15/4 | 0.22 |
| Total MME during hospitalization [median (IQR)] | 14(25) | 16(17) | 12.8(30) | 0.42 |
| VAS at Presentation/Discharge [median (IQR)] | 7(2)/0(1) | 8(1.5)/0(0) | 7(2)/0(1) | 0.53/0.46 |
| Days to advancement to solid diet [median (IQR)] | 2(1) | 2(1) | 1(1) | 0.54 |
| Respiratory failure [n (%)] | 15(2.0) | 4(2.0) | 11(1.9) | 0.70 |
| Acute Kidney Injury [n (%)] | 58 (6.2) | 15(6.6) | 43(6.2) | 0.84 |
| Bacteremia/Sepsis [n (%)] | 48(5.2) | 11(4.8) | 37(5.3) | 0.76 |
| GI bleeding [n (%)] | 37(4.0%) | 8(3.5%) | 29(4.2%) | 0.66 |
| Delirium [n (%)] | 48(5.2) | 8(3.5%) | 40(5.8%) | 0.18 |
| Discharge to rehabilitation centers [n (%)] | 27 (3) | 8(4.9) | 18(4.0) | 0.60 |

n(%): Frequency(Percentage), IQR: Interquartile Range

### Regression Analysis

The results of unadjusted univariate regression analysis performed to assess the relationship between individual outcomes of AP (dependent variable) with residence in areas with low vs. normal food geospatial access is summarized in **Table 3**. We observed no significant association between food access and AP-related mortality (*p=* 0.40) or LOS (*p=*0.99). The severity of AP, as evaluated by the modified Atlanta Criteria, demonstrated a significant association that persisted even after adjustments for demographic factors (age, Charlson Comorbidity Index (CCI), ethnoracial distribution, sex) and health care behavior variables (active alcohol and tobacco use) [OR: 1.88 (95%CI: 1.21-2.92); p = 0.005; **Table 4** and **Figure 1B**]. However, BISAP score, following adjustments for demographic and healthcare behavior variables, did not show a significant association with food access [1.07 (95%CI: 0.93-1.22); p= 0.33]. Also, there was a significant association between 30-day readmission related to AP and food access [OR: 1.78 (95%CI: 1.11-2.86); p= 0.02; **Table 4** and **Figure 1C**], and this association persisted even after adjusting for the aforementioned covariates.

**Table 3.**
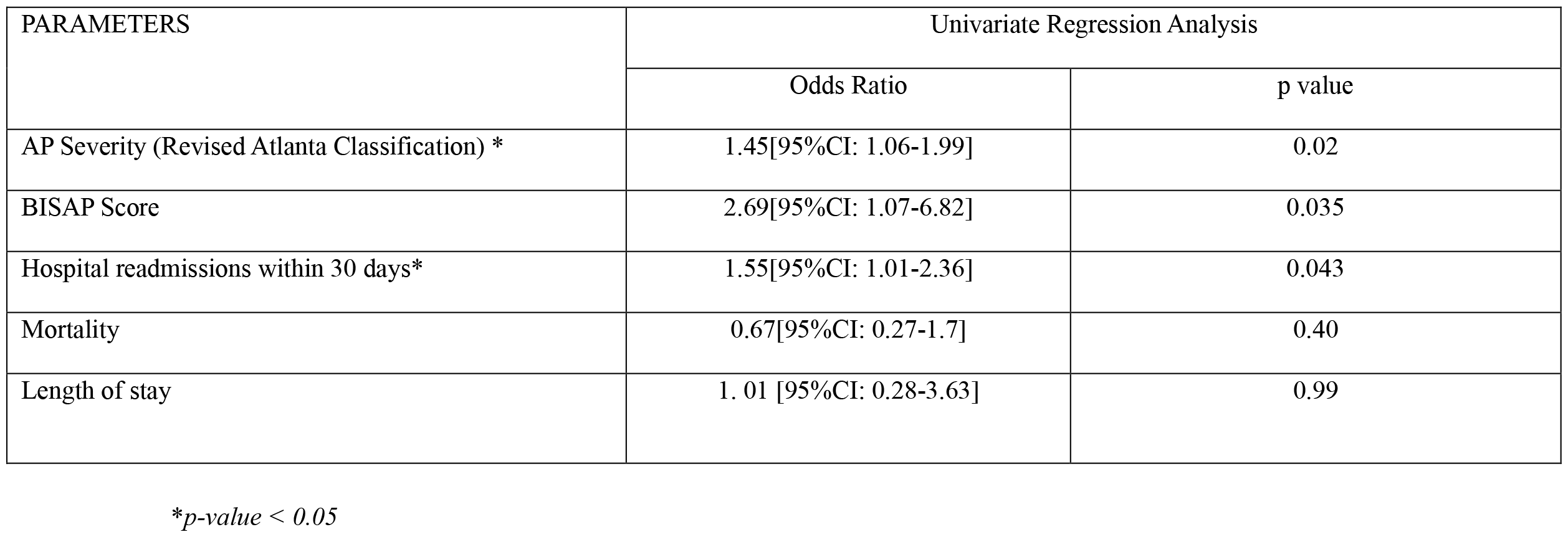
Unadjusted Odds ratios for the association of food access with acute pancreatitis outcomes:

| PARAMETERS | Univariate Regression Analysis |  |
| --- | --- | --- |
|  | Odds Ratio | p value |
| AP Severity (Revised Atlanta Classification) * | 1.45[95%CI: 1.06-1.99] | 0.02 |
| BISAP Score | 2.69[95%CI: 1.07-6.82] | 0.035 |
| Hospital readmissions within 30 days* | 1.55[95%CI: 1.01-2.36] | 0.043 |
| Mortality | 0.67[95%CI: 0.27-1.7] | 0.40 |
| Length of stay | 1. 01 [95%CI: 0.28-3.63] | 0.99 |

\**p-value* < 0.05

**Table 4.** Multivariable logistic regression analysis investigating association of acute pancreatitis outcomes with food access.

| PARAMETERS | Atlanta Classification of Severity** |  | BISAP Score |  | Hospital readmissions within 30 days** |  |
| --- | --- | --- | --- | --- | --- | --- |
|  | Odds Ratio | p value | Odds Ratio | p value | Odds Ratio | P value |
| Age | 1.01[95%CI: 0.99-1.02] | 0.24 | 1.02[95%CI: 1.01-1.03] | <0.001 | 0.97[95%CI: 0.95-0.98] | <0.001 |
| Sex (Male as reference) | 0.57[95%CI: 0.40-0.81] | 0.002 | 0.89[95%CI: 0.79-1.00] | 0.06 | 0.70 [95%CI: 0.47-1.01] | 0.06 |
| CCI | 1.02[95%CI: 0.92-1.12] | 0.73 | 1.06[95%CI: 1.03-1.10] | <0.001 | 1.18[95%CI: 1.06-1.31] | 0.002 |
| Food access (Normal access as reference) | 1.88 [95%CI:1.21-2.92] | 0.005 | 1.07[95%CI: 0.93-1.22] | 0.33 | 1.78[95%CI: 1.11-2.86] | 0.02 |
| Ethnoracial factors (White race as reference) | 1.20[95%CI: 0.81-1.80] | 0.36 | 1.02[95%CI: 0.90-1.16] | 0.75 | 0.99[95%CI: 0.65-1.51] | 0.98 |
| Active Alcohol use | 0.75[95%CI: 0.52-1.07] | 0.11 | 0.96[95%CI: 0.85-1.08] | 0.50 | 0.96[95%CI: 0.65-1.42] | 0.82 |
| Active Tobacco use | 0.91[95%CI: 0.62-1.35] | 0.65 | 0.91[95%CI: 0.80-1.04] | 0.16 | 1.21[95%CI: 0.80-1.83] | 0.36 |

\*\*Adjusting demographic variables (age, CCI, ethnoracial distribution, sex) & health care behaviors (active alcohol and tobacco use).

## DISCUSSION

The increasing prevalence of AP, as reflected in the growing rates of hospitalizations and healthcare costs, emphasizes the critical need to investigate the impact of interconnected social factors, including nutrition, healthcare behaviors, and socio-economic conditions, on the outcomes in AP. This study aims to assess how societal structures, particularly food access, may contribute to health disparities and impact the outcomes of AP patients.^27, 28^ While our primary focus is on food access, it’s crucial to recognize that food insecurity is a broader concept which includes insufficient access to food and encompasses economic and social dimensions. There is often an overlap but addressing one aspect may not fully resolve the other, and comprehensive strategies are needed to improve both food access and overall food security in a community.

Our observational study enrolled 772 patients from Massachusetts, with a substantial number (74.4%) facing low food access. We observed a higher prevalence of moderately severe and severe AP among individuals residing in regions with low food access, a significance that persisted even after adjusting for confounding factors such as demographics and health behaviors. Patient’s pain levels lacked significant association with low food access, with no differences in mortality rates or LOS between patients residing in regions with normal as compared to low food access. However, there was a significant and persistent association with 30-day readmissions and food access despite accounting for demographic factors and health behaviors. This suggests that low food access and inadequate nutrition may impact the post-discharge recovery phase and healing of AP, influencing the likelihood of readmissions. While our study concentrates on food access, the exploration of food security falls outside its scope. Among cystic fibrosis patients, both food access and food insecurity independently had a significant impact on nutritional outcomes.^3^ In patients with metabolic dysfunction-associated steatotic liver disease, food insecurity was associated with a higher incidence of fibrosis, mortality, and healthcare utilization.^4^

Despite a wealth of literature on nutrition, lifestyle, and socio-economic determinants suggesting a plausible association, direct evidence linking geospatial food access to AP outcomes and dietary patterns are lacking. Inadequate access to nutritious food may contribute to malnutrition—an established risk factor for pancreatitis.^29^ In other diseases, prior studies have also shown an association between poorer dietary choices with residence in “food swamps/deserts.” ^30, 31^ Thus, environmentally-linked dietary patterns can be hypothesized to be associated with high consumption of saturated fats especially in the context of an inflammatory condition like AP. The impact of dietary factors such as saturated fats, dietary fiber, etc. has been reported in Iowa’s women health study as well as the multiethnic cohort led by Setiawan. ^32, 33^ Both the groups observed a significantly higher risk of gallstone related AP was associated with diet rich in saturated fat and cholesterol (*e*.*g*., eggs and red meat) whereas vitamin D, milk, and fruits were associated with a reduced risk of AP. Setiawan and colleagues also noted that dietary fiber, which is known to modulate microbiome and mucosal permeability, had inverse association with severity of pancreatitis. ^33^ Hence in our study, higher rates of moderately severe and severe AP in patients residing in low food access areas may be postulated to be due to various nutritional factors including, malnutrition and diet high in saturated fats and low in fiber. Other unknown factors such as sarcopenia or preexisting pancreatic steatosis that may be related to low food access may also contribute.

Our study is unique as it is the first to investigate geospatial association between food access and outcomes related to AP. The study accessed discrete patient location which otherwise would not have been possible in large sized unidentified databases and our investigation is further strengthened by well-annotated clinical data which was meticulously gathered through manual reviews of EHR of patients with AP. Besides high diagnostic certainty, our study is notable for conservative analysis and models adjusted for potential confounding factors such as socio-demographic and health care behavior variables (smoking and alcohol consumption), which may influence the outcomes of AP. A retrospective design, although precludes the inclusion of patient-reported outcomes (including dietary patterns, preferences, food insecurity, etc.), it limits “Hawthorne effect” in patients and health-care providers thus reflecting real-time care of AP patients. ^34^

Our study is limited by the paucity of under-represented ethnoracial patients and exclusion of patients without physical addresses who have a higher risk of adverse health outcomes.^35, 36^ Single center study design at a tertiary care center also predisposes the study to referral bias. We also acknowledge the limitation of “ecological bias” in this study which may originate from attributing community or aggregate level characteristics to individuals or vice versa, as this may lead to false positive or negative causal inferences. ^37,38^ Thus, it may be inaccurate to presume all the individuals within a Federal Information Processing Standards (FIPS) have unhealthy diet patterns and reduce dietary patterns to solely food source access.

In conclusion, geospatial analyses offers promising avenues to study the impact of food access on AP outcomes. The findings of this study suggest a higher disease severity and a significant association of thirty-day readmission with disparities of food access. A causal inference of the association between food access, lifestyle, sociodemographic and other clinical outcomes in AP warrants further prospective investigation.

## Data Availability

All data produced in the present study are available upon reasonable request to the authors

---

**Supplement 1:** Strengthening the Reporting of Observational Studies in Epidemiology (STROBE) guidelines.

|  | Item<br>No | Recommendation | Page<br>No |
| --- | --- | --- | --- |
| Title and abstract | 1 | (a) Indicate the study’s design with a commonly used term in the title or the abstract | 2 |
|  |  | (b) Provide in the abstract an informative and balanced summary of what was done and what was found | 2 |
| Introduction |  |  |  |
| Background | 2 | Explain the scientific background and rationale for the investigation being reported | 3 |
| Objectives | 3 | State specific objectives, including any prespecified hypotheses | 3 |
| Methods |  |  |  |
| Study design | 4 | Present key elements of study design early in the paper | 4 |
| Setting | 5 | Describe the setting, locations, and relevant dates, including periods of recruitment, exposure, follow-up, and data collection | 4 |
| Participants | 6 | (a) Cohort study—Give the eligibility criteria, and the sources and methods of selection of participants.<br>Describe methods of follow-up | 4 |
| Variables | 7 | Clearly define all outcomes, exposures, predictors, potential confounders, and effect modifiers. Give diagnostic criteria, if applicable | 5 |
| Data source/<br>measurement | 8* | For each variable of interest, give sources of data and details of methods of assessment (measurement).<br>Describe comparability of assessment methods if there is more than one group | 6 |
| Bias | 9 | Describe any efforts to address potential sources of bias | 6 |
| Study size | 10 | Explain how the study size was arrived at | - |
| Quantitative<br>variables | 11 | Explain how quantitative variables were handled in the analyses. If applicable, describe which groupings were chosen and why | 6 |
| Statistical<br>methods | 12 | (a) Describe all statistical methods, including those used to control for confounding | 6 |
|  |  | (b) Describe any methods used to examine subgroups and interactions | 6 |
|  |  | (c) Explain how missing data were addressed | - |
|  |  | (d) Cohort study—If applicable, explain how loss to follow-up was addressed | NA |
|  |  | (e) Describe any sensitivity analyses | NA |
| Results |  |  |  |
| Participants | 13 | (a) Report numbers of individuals at each stage of study—eg numbers potentially eligible, examined for eligibility, confirmed eligible, included in the study, completing follow-up, and analyzed | 6 |
|  |  | (b) Give reasons for non-participation at each stage | 6 |
|  |  | (c) Consider use of a flow diagram | - |
| Descriptive data | 14 | (a) Give characteristics of study participants (eg demographic, clinical, social) and information on exposures and potential confounders | 6 |
|  |  | (b) Indicate number of participants with missing data for each variable of interest | 6 |
|  |  | (c) Cohort study—Summarize follow-up time (eg, average and total amount) | - |
| Outcome data | 15 | Cohort study—Report numbers of outcome events or summary measures over time | 7 |
| Main results | 16 | (a) Give unadjusted estimates and, if applicable, confounder-adjusted estimates and their precision (eg, 95% confidence interval). Make clear which confounders were adjusted for and why they were included | 7 |
|  |  | (b) Report category boundaries when continuous variables were categorized | 7 |
|  |  | (c) If relevant, consider translating estimates of relative risk into absolute risk for a meaningful time period | 7 |
| Other analyses | 17 | Report other analyses done—eg analyses of subgroups and interactions, and sensitivity analyses | NA |
| Discussion |  |  |  |
| Key results | 18 | Summarise key results with reference to study objectives | 8 |
| Limitations | 19 | Discuss limitations of the study, taking into account sources of potential bias or imprecision. Discuss both direction and magnitude of any potential bias | 9 |
| Interpretation | 20 | Give a cautious overall interpretation of results considering objectives, limitations, multiplicity of analyses, results from similar studies, and other relevant evidence | 9 |
| Generalizability | 21 | Discuss the generalizability (external validity) of the study results | 9 |
| Other information |  |  |  |
| Funding | 22 | Give the source of funding and the role of the funders for the present or the original study. | 1 |

## SUPPLEMENT 2: Determination of Geospatial Food Access

### Definition for Exposure

Geospatial food access refers to the examination of how easily individuals within a given area can obtain nutritious and affordable food options. This determination relies on a combination of the Food Access Research Data and Census tracts, with urban status being assigned according to the Bureau of the Census urban area definition.

### Low-Access Tract at 1 Mile

A tract is categorized as having low access if it meets certain criteria regarding the proximity of its residents to supermarkets, supercenters, or large grocery stores. Specifically, if a tract has a population of at least 500 people, or if 33 percent of its population resides more than 1 mile away from the nearest supermarket, supercenter, or large grocery store, it is considered to have low access. This distance threshold is particularly relevant in urban areas where residents may rely heavily on nearby food retail establishments for their grocery needs. The determination of low-access tracts relies on data sourced from the Food Access Research Atlas. This dataset provides a comprehensive directory of supermarkets, supercenters, and large grocery stores throughout the United States. This directory is compiled from multiple sources, including the 2019 STARS directory, which identifies stores authorized to accept SNAP (Supplemental Nutrition Assistance Program) benefits, and the 2019 Trade Dimensions TDLinx directory, which provides detailed information on various retail locations.

By merging these datasets, the analysis ensures a thorough representation of the spatial distribution of food retail establishments. This enabled identifying areas where access to healthy food options may be limited.

## REFERENCES

1. FAO, IFAD, UNICEF, WFP and WHO. 2018. The State of Food Security and Nutrition in the World 2018. Building climate resilience for food security and nutrition. Rome, FAO. https://www.fao.org/3/I9553EN/i9553en.pdf(Last Accessed on January 31, 2024).

2. Food Insecurity And Health Outcomes. Health Affairs 2015;34:1830–1839.

3. Bailey J, Baker E, Schechter MS, et al. Food insecurity screening and local food access: Contributions to nutritional outcomes among children and adults with cystic fibrosis in the United States. J Cyst Fibros 2023.

4. Kardashian A, Dodge JL, Terrault NA. Food Insecurity is Associated With Mortality Among U.S. Adults With Nonalcoholic Fatty Liver Disease and Advanced Fibrosis. Clin Gastroenterol Hepatol 2022;20:2790–2799 e4.

5. World Health O. A conceptual framework for action on the social determinants of health. Geneva: World Health Organization, 2010.

6. Braveman P, Gottlieb L. The social determinants of health: it’s time to consider the causes of the causes. Public Health Rep 2014;129 Suppl 2:19–31.

7. Forsmark CE, Vege SS, Wilcox CM. Acute Pancreatitis. N Engl J Med 2016;375:1972–1981.

8. Peery AF, Crockett SD, Murphy CC, et al. Burden and Cost of Gastrointestinal, Liver, and Pancreatic Diseases in the United States: Update 2021. Gastroenterology 2022;162:621–644.

9. Iannuzzi JP, King JA, Leong JH, et al. Global Incidence of Acute Pancreatitis Is Increasing Over Time: A Systematic Review and Meta-Analysis. Gastroenterology 2022;162:122–134.

10. Roberts SE, Akbari A, Thorne K, et al. The incidence of acute pancreatitis: impact of social deprivation, alcohol consumption, seasonal and demographic factors. Aliment Pharmacol Ther 2013;38:539–48.

11. Petrov MS, Yadav D. Global epidemiology and holistic prevention of pancreatitis. Nat Rev Gastroenterol Hepatol 2019;16:175–184.

12. Li CL, Jiang M, Pan CQ, et al. The global, regional, and national burden of acute pancreatitis in 204 countries and territories, 1990-2019. BMC Gastroenterol 2021;21:332.

13. Lankisch PG, Apte M, Banks PA. Acute pancreatitis. Lancet 2015;386:85–96.

14. Navina S, Acharya C, DeLany JP, et al. Lipotoxicity causes multisystem organ failure and exacerbates acute pancreatitis in obesity. Sci Transl Med 2011;3:107ra110.

15. Fagenholz PJ, Fernandez-del Castillo C, Harris NS, et al. Direct medical costs of acute pancreatitis hospitalizations in the United States. Pancreas 2007;35:302–7.

16. Wang Z, Guo M, Li J, et al. Composition and functional profiles of gut microbiota reflect the treatment stage, severity, and etiology of acute pancreatitis. Microbiol Spectr 2023;11:e0082923.

17. von Elm E, Altman DG, Egger M, et al. Strengthening the Reporting of Observational Studies in Epidemiology (STROBE) statement: guidelines for reporting observational studies. Bmj 2007;335:806–8.

18. International Classification of Diseases, Tenth Revision, Clinical Modification (ICD-10-CM); [updated 2017 Aug 18; reviewed 2017 Aug 18; cited 2017 Oct 11]. Available from: http://www.cdc.gov/nchs/icd/icd10cm.html (Last Accessed on January 31, 2024).

19. International Classification of Diseases NR, Clinical Modification (ICD-9-CM); [updated 2010 Sep 21; reviewed 2010 Sep 21; cited 2010 Nov 23]. Available from: http://www.cdc.gov/nchs/icd/icd9cm.htm. (Accessed on January 31, 2024).

20. Banks PA, Bollen TL, Dervenis C, et al. Classification of acute pancreatitis--2012: revision of the Atlanta classification and definitions by international consensus. Gut 2013;62:102–11.

21. Economic Research Service (ERS) USDoAUFARA, https://www.ers.usda.gov/data-products/food-access-research-atlas/. (Last Accessed on January 31,2024).

22. Institute of Medicine and National Research Council. 2013. Supplemental Nutrition Assistance Program: Examining the Evidence to Define Benefit Adequacy. Washington DTNAPhd. (Last Accessed on January 31,2024).

23. https://nielseniq.com/global/en/solutions/tdlinx/. (Last Accessed on January 31,2024).

24. McHenry N, Shah I, Ahmed A, et al. Racial Variations in Pain Management and Outcomes in Hospitalized Patients With Acute Pancreatitis. Pancreas 2022;51:1248–1250.

25. Anderson KL, Shah I, Tintara S, et al. Evaluating the Clinical Characteristics and Outcomes of Idiopathic Acute Pancreatitis: Comparison With Nonidiopathic Acute Pancreatitis Over a 10-Year Period. Pancreas 2022;51:1167–1170.

26. Charlson ME, Pompei P, Ales KL, et al. A new method of classifying prognostic comorbidity in longitudinal studies: development and validation. J Chronic Dis 1987;40:373–83.

27. Brindise E, Elkhatib I, Kuruvilla A, et al. Temporal Trends in Incidence and Outcomes of Acute Pancreatitis in Hospitalized Patients in the United States From 2002 to 2013. Pancreas 2019;48:169–175.

28. Gapp J, Hall AG, Walters RW, et al. Trends and Outcomes of Hospitalizations Related to Acute Pancreatitis: Epidemiology From 2001 to 2014 in the United States. Pancreas 2019;48:548–554.

29. Mao X, Huang C, Wang Y, et al. Association between Dietary Habits and Pancreatitis among Individuals of European Ancestry: A Two-Sample Mendelian Randomization Study. Nutrients 2023;15.

30. Cooksey Stowers K, Jiang Q, Atoloye A, et al. Racial Differences in Perceived Food Swamp and Food Desert Exposure and Disparities in Self-Reported Dietary Habits. Int J Environ Res Public Health 2020;17.

31. Berkowitz SA, Karter AJ, Corbie-Smith G, et al. Food Insecurity, Food “Deserts,” and Glycemic Control in Patients With Diabetes: A Longitudinal Analysis. Diabetes Care 2018;41:1188–1195.

32. Prizment AE, Jensen EH, Hopper AM, et al. Risk factors for pancreatitis in older women: the Iowa Women’s Health Study. Ann Epidemiol 2015;25:544–8.

33. Setiawan VW, Pandol SJ, Porcel J, et al. Dietary Factors Reduce Risk of Acute Pancreatitis in a Large Multiethnic Cohort. Clin Gastroenterol Hepatol 2017;15:257–265 e3.

34. Sedgwick P, Greenwood N. Understanding the Hawthorne effect. BMJ 2015;351:h4672.

35. Fazel S, Geddes JR, Kushel M. The health of homeless people in high-income countries: descriptive epidemiology, health consequences, and clinical and policy recommendations. Lancet 2014;384:1529–40.

36. Doran KM, Boyer AP, Raven MC. Health Care for People Experiencing Homelessness-What Outcomes Matter? JAMA Netw Open 2021;4:e213837.

37. Greenland S, Morgenstern H. Ecological bias, confounding, and effect modification. Int J Epidemiol 1989;18:269–74.

38. Loney T, Nagelkerke NJ. The individualistic fallacy, ecological studies and instrumental variables: a causal interpretation. Emerg Themes Epidemiol 2014;11:18.

